# Naturally-acquired Immunity Dynamics against SARS-CoV-2 in Children and Adolescents

**DOI:** 10.1101/2022.06.20.22276650

**Authors:** Tal Patalon, Yaki Saciuk, Hanit Ohayon Hadad, Galit Perez, Asaf Peretz, Amir Ben-Tov, Sivan Gazit

## Abstract

**Objectives:** There are paucity of studies examining naturally acquired immunity against SARS-CoV-2 in children and adolescents, though they are generally the last group to be afforded the vaccine, and a significant portion of them are still unvaccinated. This study examined the duration of protection conferred by a previous SARS-CoV-2 infection amongst children and adolescents.

**Design:** A retrospective study, applying two complementary approaches: a matched test-negative case control design and a retrospective cohort design.

**Setting:** Nationally centralized database of Maccabi Healthcare Services, an Israeli national health fund that covers 2.5 million people.

**Participants:** The study population included between 293,743 and 458,959 individuals (depending on the model), 5-18 years of age, who were unvaccinated SARS-CoV-2 naïve persons or unvaccinated convalescent patients.

**Main outcomes and measures:** Analyses focused on the period of July 1 to December 13, 2021, a Delta-dominant period in Israel. We evaluated three SARS-CoV-2-related outcomes: (1) documented PCR confirmed infection or reinfection, (2) COVID-19 and (3) severe COVID-19.

**Results:** Overall, children and adolescents who were previously infected acquired durable protection against reinfection (symptomatic or not) with SARS-CoV-2 for at least 18 months. Importantly, no COVID-19 related deaths were recorded in either the SARS-CoV-2 naïve group or the previously infected group. Effectiveness of naturally-acquired immunity against a recurrent infection reached 89.2% (95% CI: 84.7%-92.4%) three to six months after first infection, mildly declining to 82.5% (95% CI, 79.1%-85.3%) nine months to one year after infection, then remaining rather steady for children and adolescents for up to 18 months, with a slight non-significant waning trend. Additionally, we found that ages 5-11 exhibited no significant waning of naturally acquired protection throughout the outcome period, whereas waning protection in the 12-18 age group was more prominent, but still mild.

**Conclusions:** Children and adolescents who were previously infected with SARS-CoV-2 remain protected against reinfection to a high degree for 18 months. Policy decision makers should consider when and if convalescent children and adolescents should be vaccinated. Nonetheless, further research is needed to examine naturally acquired immunity against emerging variants, including the Omicron.

## Introduction

The duration of protection conferred by a prior COVID-19 episode against reinfection with SARS-CoV-2 has been studied, though not yet conclusively determined. The implications of understanding the duration of naturally acquired immunity are vast, such as the necessity and timing of actively pursuing hybrid immunity (i.e., vaccinating convalescent patients), mandating self-quarantine after exposure of previously infected individuals and more.

Two main challenges complicate the research of naturally acquired immunity; the first is the lack of evidence-based, long-term correlate of protection^1^. The second is the difficulty in defining reinfection as opposed to prolonged viral shedding^2^. In an ideal scenario, reinfection would be determined by two distinct clinical episodes with two different sequenced viral genomes. Nonetheless, as sequencing is seldom performed in routine population tests, these strict criteria are impossible to apply in real-world data research.

Therefore, different criteria based on more widely-available data have been suggested.^3^ The predominant one have been published by The Centers for Disease Control and Prevention’s (CDC), where guidelines refer to two positive SARS-CoV-2 polymerase chain reaction (PCR) test results at least 90 days apart.^4^

In adults, studies have demonstrated that naturally acquired immunity^5,6^ appears to provide protection against reinfection for between at least 7 and 13 months,^7^ though protection was age-dependent and lower for those 65 years or older.^8^ Furthermore, while correlating viral loads to infectiousness, studies have shown that reinfections are less infectious than primary infections, and possibly less so than breakthrough infections (i.e., infections in fully vaccinated individuals).^9^ Correspondingly, studies have shown that in adults naturally acquired immunity lasts longer and confers a stronger protection relatively to vaccine-induced immunity.^10^

However, there are paucity of studies examining naturally acquired immunity in children and adolescents, though they are generally the last group to be afforded the vaccine, and a significant portion of them are still unvaccinated. Israel, an early adopter of the SARS-CoV-2 vaccine in adults, launched a vaccination campaign for adolescents on June, 2021,^11^ (shortly after the BioNTech/Pfizer mRNA BNT162b2 vaccine was approved for adolescents by the United States Food and Drug Administration (FDA) and the European Medicines Agency in May 2021),^12,13^ and for children ages 5-11 on November, 2021. Two studies that investigated the rates of recurrent infection in children reached similar conclusion; a study from California^14^ found lower reinfection rates in children, as compared to adults, and similarly, a study from England^15^ found a lower risk of reinfection in children. Nonetheless, the questions of waning of infection-induced-immunity, or the durability of protection, have yet to be answered.

To this end, we conducted a retrospective study evaluating the duration of protection conferred by a previous SARS-CoV-2 infection amongst children and adolescents aged 5 to 18, leveraging data from Maccabi Healthcare Services (MHS), Israel’s second largest Health Maintenance Organization that covers 2.5 million members. Relying on 21 months of data, from March 1, 2020 to December 13, 2021, this is the longest study published on naturally acquired immunity in this age group.

## Methods

### Definitions and nomenclature

Individuals with a documented SARS-CoV-2 infection, determined by a previous positive PCR test, were referred to as “previously infected”, “convalescent” or “recovered” individuals. SARS-CoV-2 reinfection (see under ***Measured Outcomes***) was defined as a positive PCR test during the outcome period, confirmed by a PCR test, regardless of the existence of symptoms. Individuals who had a documented PCR test *and* at least one documented COVID-19-related-symptom were considered as having a “symptomatic infection” or “COVID-19”. Of those, hospitalized patients were referred to as incurring “COVID-19-related hospitalization”. Lastly, the protection conferred by a previous infection was denoted as “naturally acquired immunity”, or “infection-induced immunity”. SARS-CoV-2 naïve individuals indicated persons who have never had a record of a positive SARS-CoV-2 test.

### Reinfection with SARS-CoV-2

In investigating large datasets of COVID-19 information, a challenge exists in defining SARS-CoV-2 reinfection (or a ‘failure’ of infection-induced immunity) as opposed to prolonged viral shedding^2^. This study will adhere to the Centers for Disease Control and Prevention’s (CDC) guidelines, which define a reinfection as two positive SARS-CoV-2 polymerase chain reaction (PCR) test results at least 90 days apart.^16^

### Data sources

MHS is a large not-for-profit health fund in Israel, covering 26.7% of the population which constitute a representative sample of the Israeli population. Healthcare services are provided throughout the country, through over 150 local branches (or clinics), situated within a walking or short driving distance from each member’s registered address. MHS has maintained a centralized database of Electronic Medical Records (EMRs) for three decades, where less than 1% disengagement rate among its members has allowed for a comprehensive longitudinal medical follow-up. The centralized database includes extensive demographic data, clinical measurements and evaluations, outpatient and hospital diagnoses and procedures, medications dispensed, imaging performed and comprehensive laboratory data from a single central laboratory.

### Data extraction

Individual-level demographic data of the study population included age, sex, socioeconomic status (SES), one’s MHS branch and a coded geographical statistical area (GSA, assigned by Israel’s National Bureau of Statistics, corresponds to neighborhoods and is the smallest geostatistical unit of the Israeli census). The SES is measured on a scale from 1 (lowest) to 10, and the index is calculated based on several parameters, including household income, educational qualifications, household crowding and car ownership. Data about one’s medical history included the last documented Body Mass Index (BMI) (where obesity was defined as BMI ≥ 30) and information about chronic diseases from MHS’ automated registries, including cardiovascular diseases^17^, hypertension^18^, diabetes^19^, inflammatory bowel diseases, chronic kidney disease^20^, chronic obstructive pulmonary disease, immunocompromised conditions, and cancer from the National Cancer Registry^21^.

COVID-19-related information was also captured, comprising of dates of SARS-CoV-2 vaccination and results of any PCR test, including one performed outside of MHS, given that all such tests are recorded centrally. Records of COVID-19-related symptoms were extracted from members’ EMRs, where they were recorded by the primary care physician or a certified nurse who conducted in-person or phone visits with infected individual. Finally, data on COVID-19-related hospitalizations were retrieved as well, and COVID-19-related mortality was screened for.

### Study population

The study population included all MHS members aged 5-18 years who were unvaccinated SARS-CoV-2 naïve persons or unvaccinated convalescent patients. Age and immunity status (including vaccination and previous infection) were assessed on the day of inclusion (the latter different between the two study designs, see: ***design and statistical analysis***), and for convalescent patients, the prior infection must have occurred at least 90 days prior to the inclusion date, in order to capture reinfections (as opposed to prolonged viral shedding) by following the 90-day guideline of the CDC (see: ***reinfection with SARS-CoV-2***). We excluded individuals who received any dose of the SARS-CoV-2 vaccine prior to their inclusion in the study and persons with a possibly incomplete COVID-19-related medical history during the pandemic, i.e. those who joined MHS after March 2020. Additionally, we excluded any individual who had already been re-infected by the start of the outcome period, in order to avoid potential bias by aggregating twice- and thrice-COVID-19 patients.

### Study design and statistical analysis

#### Measured outcomes

We evaluated three SARS-CoV-2-related outcomes: documented RT-PCR confirmed SARS-CoV-2 infection or reinfection, COVID-19 and severe COVID-19, defined as COVID-19-related hospitalization or death. Outcomes were evaluated during the follow-up period of July 1 to December 13, 2021, a Delta-dominant period in Israel.^22^

#### Comparing effectiveness over time

Prior SARS-CoV-2 vaccination studies demonstrated a time-dependent increase and then decrease in the level of protection conferred by the vaccine.^23–27^ This analysis explored whether a reduction in *infection*-conferred protection exists as well, by using a similar metric of comparison to that of previous *vaccination* studies. Therefore, we performed a stratified analysis by time-since-infection into equal 90-day intervals (3-6, 6-9, 9-12, 12-15, 15-18 months since previous infection). The rationale was that if no waning of naturally acquired immunity existed, reinfections would not depend on time-since-previous-infection. However, if waning existed, protection conferred by previous infection would gradually decrease with time.

We conducted two complementary approaches, both previously used to evaluate effectiveness of protection against SARS-CoV-2. Our primary analysis included a matched test-negative case control design, whereas the secondary one compared infection rates applying a retrospective cohort design.

#### Primary analysis – test-negative case-control design

For our primary analysis, we used a test-negative matched case-control design, widely used in COVID-19 studies owing to its ability to control for bias stemming from misclassification of untested persons, healthcare-seeking- and testing-related behaviors.^23,27–29^ We defined cases as (1) individuals with a SARS-CoV-2-positive PCR test during the study period, (2) individuals with a symptomatic SARS-CoV-2 infection, or (3) persons with COVID-19-related hospitalization or death, where cases were defined separately for each outcome. Eligible controls were individuals with a negative PCR test result during the outcome period.

We performed a 1:1 matching based on sex, GSA and week of testing (to account for potential time-varying risk within the outcome period). The first positive PCR test and the first negative PCR test were the only tests included for each case and control, respectively. All negative PCRs for cases were excluded from the study, therefore an MHS member was either a case or a control, not both.^25,26^

The analysis sought to estimate the odds of a positive test at the different time points following infection, compared to SARS-CoV-2 naïve individuals. Therefore, we stratified the analysis for each time-since-infection interval, where in each stratum we included only participants that met that specific 90-day interval or the reference group of SARS-CoV-2 naïve individuals (consequently, the number of matched pairs varied between time points).^25^ One’s ‘immunity-status’ (time-since-previous-infection) was determined by the date of the PCR test.

Conditional logistic regression models were fit to the data, accounting for the matching. The effectiveness of a previous infection at each time point, compared to SARS-CoV-2 naïve individuals, was calculated as 100%*[1-(Odds Ratio)] for each of the 3-month-since-infection intervals.

As an additional analysis, we sought to evaluate whether the pattern of naturally acquired immunity against reinfection varies with age. Therefore, we measured the interaction of two groups – 5-11 and 12-18 years – with each time-since-first-infection interval. These age groups were chosen as they correspond to the vaccination rollout in Israel, where the younger age group first became eligible months after the older one. Though our analysis focuses on unvaccinated children, we presumed some behavioral influences relating to the post-vaccination behavior of their respective peers of the same age group, which might lead to varying exposures.

#### Secondary analysis – a retrospective cohort design

In our secondary analysis, we compared the incidence rates of the three outcomes (SARS-CoV-2 infection, COVID-19 disease, and severe diseases) during the follow-up period between recovered individuals (with different times since recovery) and SARS-CoV-2 naïve ones, where the latter were the reference group. These cohorts were dynamic, as for example, participants who were part of the 3-6-months-since-infection group on July 1, 2021, left it on October, and joined the 6-9-months-after-infection group, as long as no measured outcome occurred during these months (namely SARS-CoV-2 infection, disease or severe disease) as well as no censoring event (i.e. vaccination or leaving MHS). (**Figure S1**). Therefore, individuals could contribute follow-up days to multiple groups, and move between them, according to their immunity status on each follow-up day. For each time-since-infection group we calculated the rate of each outcome per person days at risk. Similarly to the primary model, we reran the analysis while measuring the interaction of time-since-firs-infection with a younger (5-11) and older (12-18) age group.

A Poisson regression model was fit to the data, adjusting for (1) a categorical variable consisting of the number of previous PCR tests performed by each individual since the start of the pandemic (March 2020) until the start of the outcome period (as a proxy for SARS-CoV-2-related healthcare-seeking behavior, differentiating *behavior* from outcome),^30,31^ and (2) a residential-specific *attack rate*, calculated for each day in which a PCR test was performed, representing the average proportion of infected MHS members belonging to a specific branch within that calendar week, divided by the overall number of participants registered to that branch (of that specific city, village or town). Adjusting for attack rate allowed us to address the time-varying and geographical-varying risk of exposure, which might influence effectiveness. These two adjustments are the Poisson’s answer to the test-negative design, where time of testing and GSA were matched.^32^

Then, we calculated the rate ratio, comparing the incidence rate of time-since-infection to that of the SARS-CoV-2 naïve participants for each time-since-infection interval. Similarly to the primary model, the effectiveness of a previous infection at each time point, compared to SARS-CoV-2 naïve individuals, was calculated as 100%*[1-(Rate Ratio)] for each of the 3-month-since-infection intervals.

Moreover, we applied an indirect standardization of rates per 100,000 person days, based on the parameters of the fitted regression analysis, yielding adjusted rates for each time-since-infection intervals, therefore projecting the rates as if all person days at risk were included in each time-since-infection interval. 95% confidence intervals were computed using the bootstrap method.

All analyses were conducted using R Studio version 3.6 with the survival package. The analysis conforms to Strengthening the Reporting of Observational Studies in Epidemiology (STROBE) Statement.

## Data Availability

According to the Israel Ministry of Health regulations, individual-level data cannot be shared openly. Specific requests for remote access to de-identified community-level data should be referred to KSM, Maccabi Healthcare Services Research and Innovation Center.

## Ethics declaration

This study was approved by the MHS (Maccabi Healthcare Services) Institutional Review Board (IRB). Due to the retrospective design of the study, informed consent was waived by the IRB, and all identifying details of the participants were removed before computational analysis.

## Code availability statement

Specific requests for remote access to the code used for data analysis should be referred to KSM, Maccabi Healthcare Services Research and Innovation Center.

## Results

### Primary analysis: naturally acquired immunity against infection

293,743 MHS members aged 5-18 years were eligible for the study and obtained at least one PCR test between July 1 and December 13, 2021. Across all test-negative analyses, 200,329 cases of infection and 32,829 cases of symptomatic infection were included, to which controls were matched. The peak incidence occurred between August and September 2021 (**Figure S2**). 50 individuals incurred a COVID-19 related hospitalization, and no COVID-19 related deaths were recorded during the study period. Baseline characteristics of participants are given in **Table 1**. Overall, the population was healthy and demographic characteristics were similar between the groups.

**Table 1.**
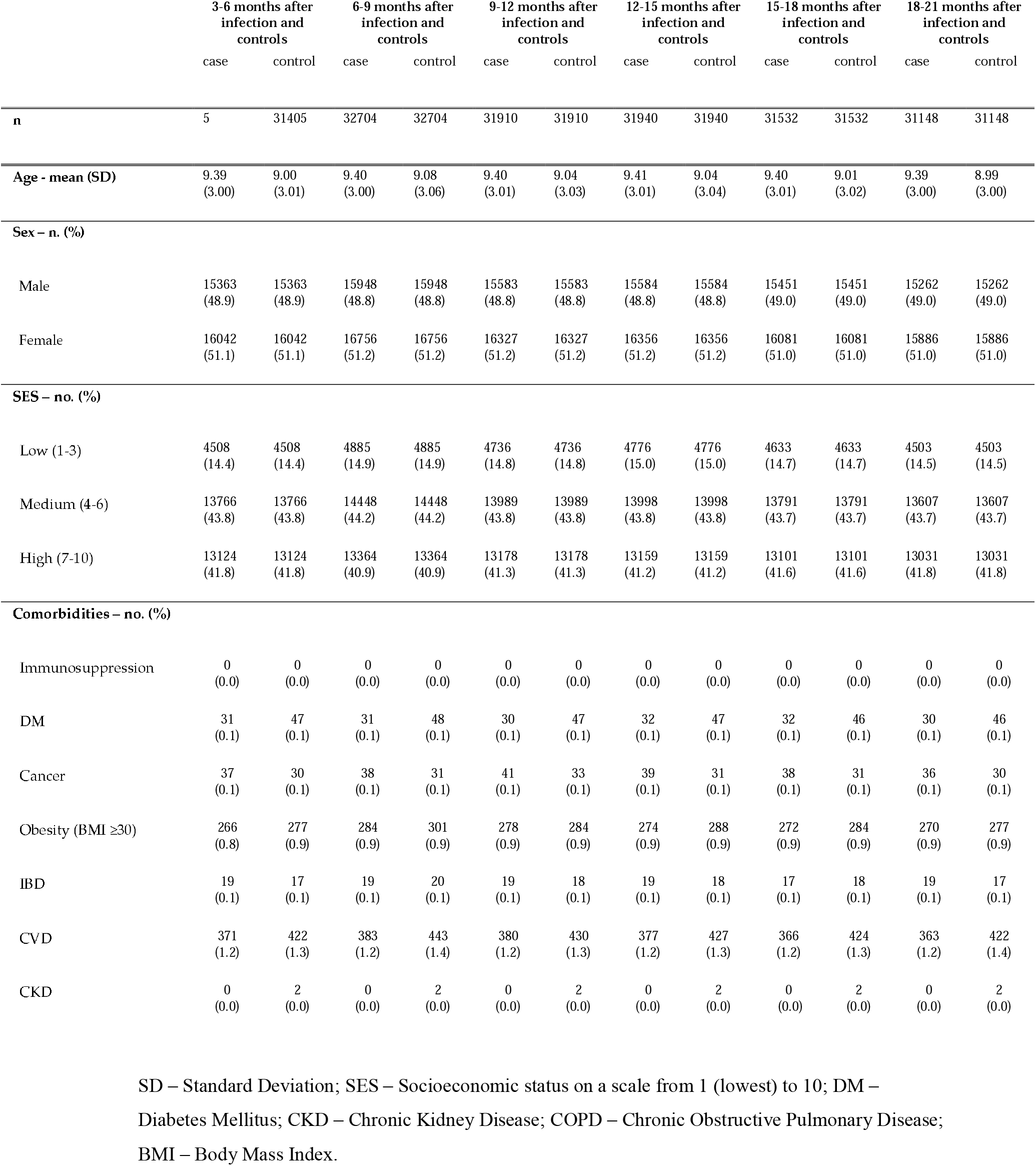
Characteristics of unvaccinated participants 5-18 years old who were tested between July 1 and December 13, 2021, by groups. Matching was based on sex, GSA and week of testing.

Estimates of effectiveness of naturally acquired immunity by time since infection are presented in **Table 2**. Effectiveness was based on the odds ratios (OR), which represented the ratio between the odds of testing positive for SARS-CoV-2 during the outcome period among SARS-CoV-2 naïve persons (the reference group) and the respective odds of infection among convalescent patients in each subsequent months-interval.

**Table 2.** Testing results at different times-since-infection and effectiveness of naturally-acquired-immunity against SARS-CoV-2 infection. Effectiveness was defined as 100%*[1-(Odds Ratio)] for each of the time-since-vaccination intervals. The reference group comprised of SARS-CoV-2 naïve individuals, where analysis applied a 1:1 matching, based on sex, GSA and week of testing.

|  | Cases (PCR positive) |  | Controls (PCR negative) |  | Effectiveness of naturally acquired immunity (%) against SARS-CoV-2 infection (95% CI) |
| --- | --- | --- | --- | --- | --- |
|  | Previously infected | SARS-CoV-2 naïve | Previously infected | SARS-CoV-2 naïve |  |
| Months after infection |  |  |  |  |  |
| 3-6 | 35 | 32974 | 315 | 32694 | 89.2(84.7, 92.4) |
| 6-9 | 215 | 33407 | 1621 | 32001 | 88.2(86.4, 89.8) |
| 9-12 | 153 | 33174 | 799 | 32528 | 82.5(79.1, 85.3) |
| 12-15 | 186 | 33160 | 828 | 32518 | 79.5(75.8, 82.6) |
| 15-18 | 93 | 33021 | 411 | 32703 | 78.4(72.8, 82.8) |
| 18-21 | 17 | 33894 | 38 | 32873 | 57.1(22.7, 76.2) |

The effectiveness against re-infection of individuals 3 to 6 months since previous infection was 89.2% (95% Confidence Interval [CI]: 84.7%-92.4%) compared to those not previously infected (**Table 2, Figure 1**). Infection-induced immunity mildly declined to 82.5% (95% CI, 79.1%-85.3%) in the 9-12-months-since-infection window. Protection of those who were infected 12 to 15 months prior and 15 to 18 months prior to the study outcome period had overall comparable odds of infection with the 9-12-since-infection group, as no statistical difference between these intervals could be concluded. Few observations after 18 months of infection led to a wide confidence interval that challenges inferences, apart from suggesting those previously infected even 21 months prior to the outcome period remain protected to some degree compared to uninfected children and adolescents.

**Figure 1.**
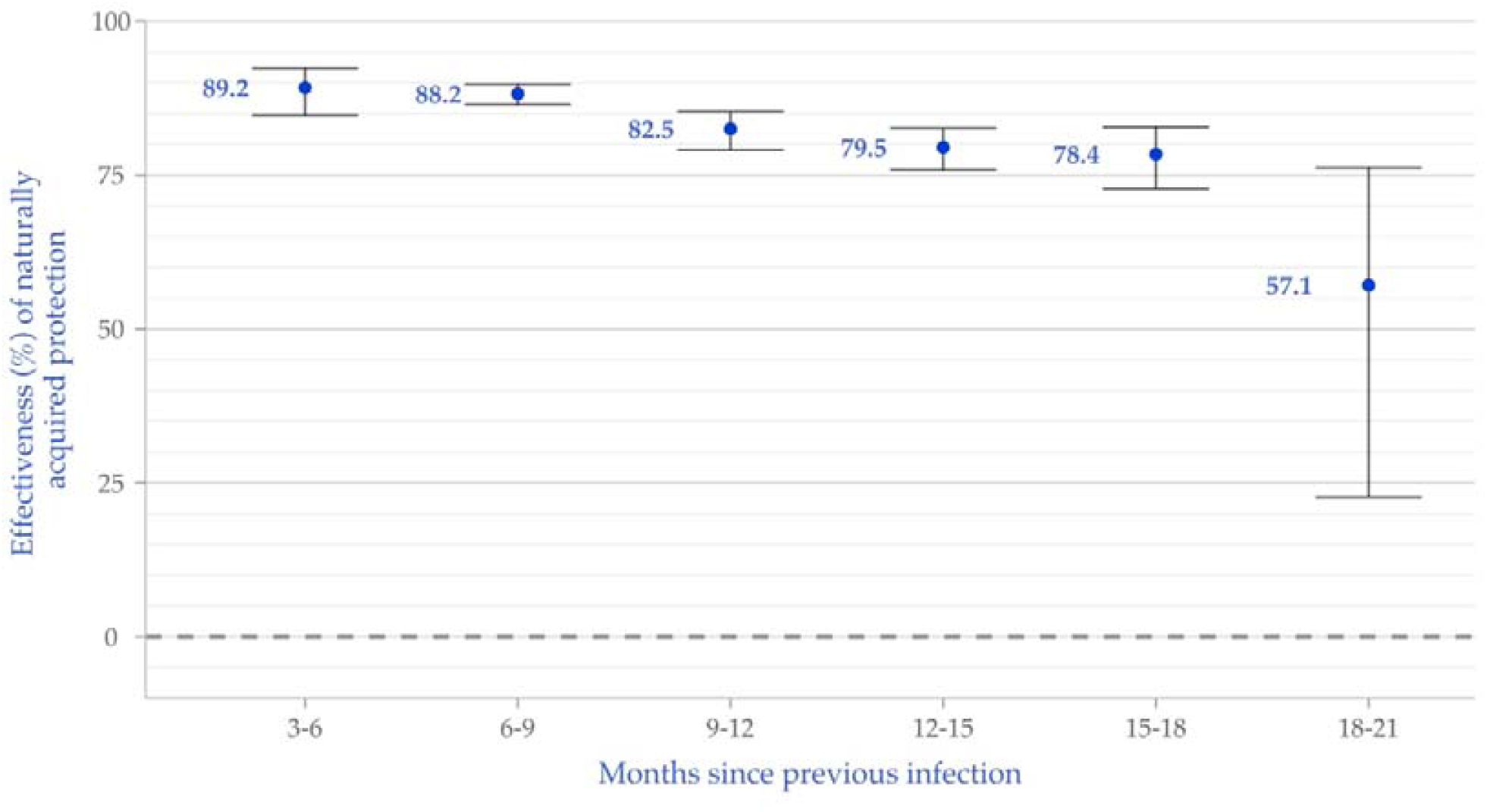
Effectiveness of naturally acquired immunity compared to SARS-CoV-2 naïve individuals against infection, by time since previous infection, based on the primary test-negative analysis. Bars represent 95% confidence intervals.

When examining the interaction with the younger and older age groups, we found that the protection conferred by previous infection was roughly similarly effective against reinfection in each 3-months interval in the younger age group of 5-11 years old, with a slight waning trend throughout 18 months. Contrastingly, there was a trend toward a more rapid waning of protection in the 12-18 years old group, though relative paucity of observations yielded wider confidence intervals (**Figure S3**). The interval of 18-21 months after previous infection was not included in the analysis due to too few observations.

### Primary analysis: naturally acquired immunity against symptomatic infections (COVID-19)

Effectiveness of a previous infection against re-infection resulting in a symptomatic disease was slightly higher than that against any re-infection (regardless of symptoms) 3-6 months after infection, at 93.6% (95% CI, 79.6%-98.0%), and presented a possibly more rapid trend of decline with time since infection, though the differences in effectiveness between the different time-since-previous-infection intervals was not statistically significant (**Table 3, Figure 2**) and protection against a symptomatic reinfection remained high. The interval of 18-21 months after previous infection was dropped in this analysis as well, due to too few observations.

**Table 3.** Testing results at different times-since-infection and effectiveness of naturally-acquired-immunity against COVID-19. Effectiveness was defined as 100%*[1-(Odds Ratio for symptomatic infection)] for each of the time-since-vaccination intervals. The reference group comprised of SARS-CoV-2 naïve individuals, where analysis applied a 1:1 matching, based on sex, GSA and week of testing.

|  | Cases (PCR positive) |  | Controls (PCR negative) |  | Effectiveness of naturally acquired immunity (%) against COVID-19 (95% CI) |
| --- | --- | --- | --- | --- | --- |
|  | Previously infected | SARS-CoV-2 naïve | Previously infected | SARS-CoV-2 naïve |  |
| Months after infection |  |  |  |  |  |
| 3-6 | 3 | 6537 | 46 | 6494 | 93.6(79.3, 98.0) |
| 6-9 | 35 | 6555 | 229 | 6361 | 85.5(79.2, 89.9) |
| 9-12 | 26 | 6540 | 114 | 6452 | 78.3(66.5, 86.0) |
| 12-15 | 33 | 6543 | 125 | 6451 | 76.3(64.5, 84.2) |
| 15-18 | 22 | 6535 | 70 | 6487 | 69.8(50.7, 81.5) |

**Figure 2.**
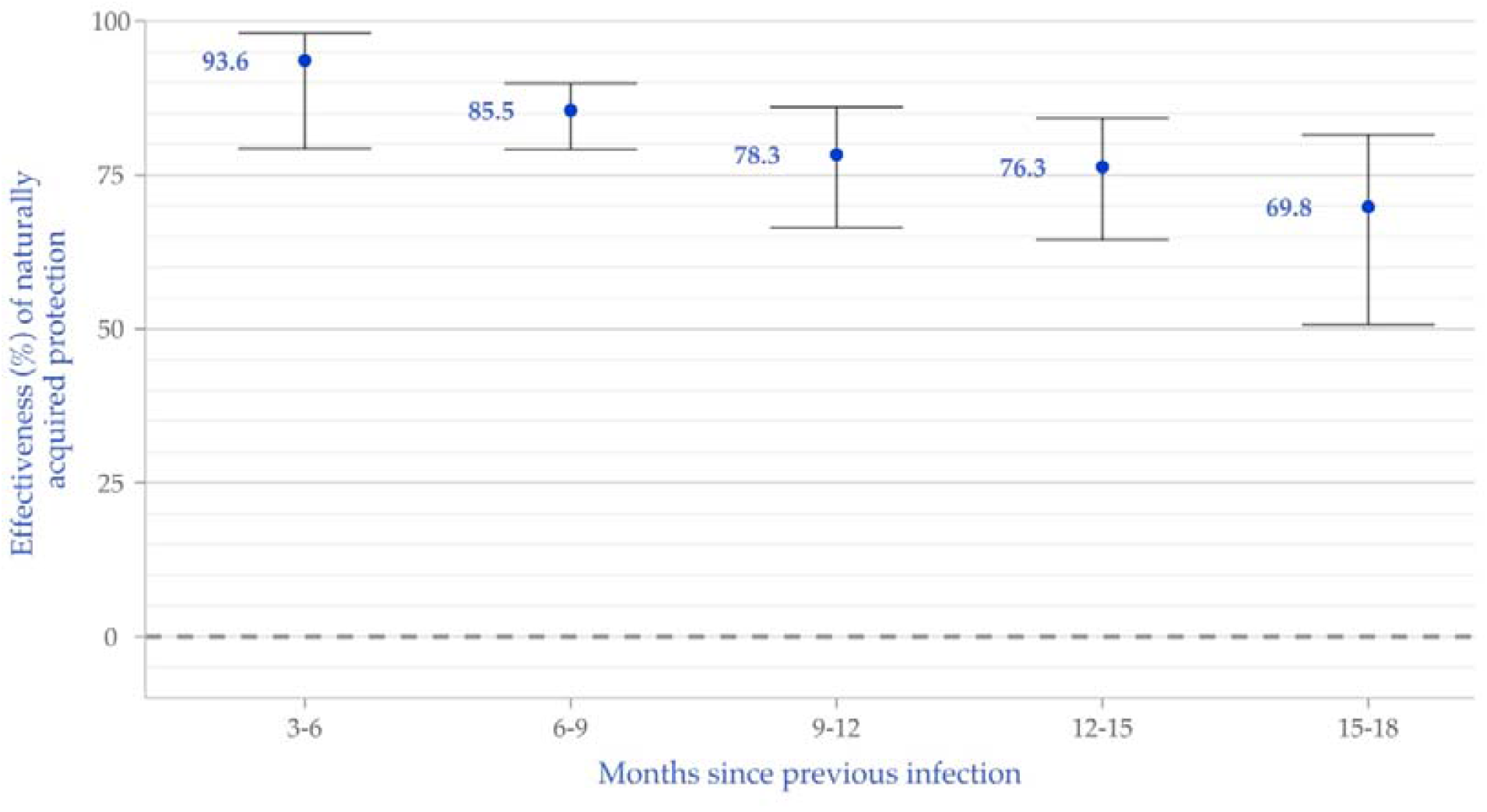
Effectiveness of naturally acquired immunity compared to SARS-CoV-2 naïve individuals against symptomatic COVID-19, by time since previous infection, based on the primary test-negative analysis. Bars represent 95% confidence intervals.

As for the outcome of severe disease, no deaths were recorded, and relatively few COVID-19-related hospitalizations were recorded; 48 in SARS-CoV-2 naïve individuals, and 2 in those previously infected. These numbers did not allow for a reliable regression analysis.

### Secondary analysis – Poisson regression model

In our secondary model, 458,959 individuals contributed 57,737,823 person-days at risk, during which 40,516 and 19,585 SARS-CoV-2 infections (or reinfections) and symptomatic infections (or reinfections) occurred, respectively. Their clinical and demographic characteristics can be seen in **Table S1**. In this model as well, the groups had overall similar demographic and clinical attributes, apart from the group of 18-21 months after infection, which had lower socioeconomic status, and higher rates of comorbidities. However, due to few observations in this group, inference of naturally acquired protection in group was limited and dropped from the analysis, therefore the possible effect of these differences was not assessed.

**Table S2** details the results of the Poisson regression analysis, where the rate ratio of re-infection for each time-since-first-infection group relative to SARS-CoV-2 naïve individuals is presented. Effectiveness, calculated as 100%*[1-(Rate Ratio)], can be seen in **Figure S4**. Additionally, **Table S2** presents the adjusted incidence rates of SARS-CoV-2 infection per 100,000 person days for each time-since-infection group, using indirect standardization. Effectiveness was overall similar to the primary test-negative analysis, though confidence intervals were larger.

When measuring the interaction of time-since-infection with age group, a pattern similar to the primary analysis emerged, where the 5-11 age group showed no significant difference in effectiveness throughout the 18 months intervals, whereas the 12-18 age group demonstrated a significant decline in effectiveness of naturally acquired protection against re-infection after 12 months (**Figure S5**).

As for protection against a symptomatic re-infection conferred by previous infection, results are similar to the primary model in this analysis as well (**Table S2, Figure S6**), and effectiveness is slightly higher against reinfection that results in a symptomatic disease - as opposed to any re-infection, regardless of symptoms. No significant waning of protection is seen between groups of different times-since-first-infection, but only a trend.

## Discussion

This study evaluated the dynamics and durability of naturally acquired immunity against reinfection with SARS-CoV-2 in children and adolescents. It is the largest real-world observational research examining this question to date, encompassing 21 months of longitudinal data, and including between 293,743 and 458,959 individuals, depending on the model of analysis.

Waning of natural immunity can be conceptualized as decreased protection against re-infection with time-since-first-infection (within the same calendrical outcome period); our aim was to examine whether the protection waned – and when. To this end, our primary analysis took a test-negative approach, and compared two groups of unvaccinated individuals: SARS-CoV-2 naïve persons and previously infected ones during the same outcome period. We further stratified the convalescent group according to time since first infection.

Overall, children and adolescents who were previously infected acquired strong and durable protection against reinfection (symptomatic or not) with SARS-CoV-2 for at least 18 months. Importantly, no COVID-19 related deaths were recorded in either the SARS-CoV-2 naïve group or the previously infected group.

Effectiveness of naturally-acquired immunity against a recurrent SARS-CoV-2 infection reached 89.2% three to six months after first infection, mildly declining to 82.5% nine months to one year after infection, then remaining rather steady for children and adolescents up to 18 months, with a slight non-significant waning trend. Those infected 18 to 21 months prior to the outcome period were still protected, though scarce data rendered it difficult to quantify. Naturally-acquired immunity against a *symptomatic* re-infection (COVID-19), conferred relatively similar protection to that against a SARS-CoV-2 infection - regardless of symptoms.

As for public health policies, the demonstrated long-term protection of naturally acquired immunity has important implications regarding the decision to vaccinate convalescent children and adolescents, and to mandate self-quarantine after exposure, affecting all biopsychosocial aspects of life and well-being of children, adolescents and their families. This should be considered in light of evidence of increasing seroprevalence (of anti-nucleocapsid (anti-N) antibodies, indicating previous infection) in children and adolescents, crossing 70% in the United States in 2022.^33^

Our findings prolongs the duration of natural immunity compared to previous publications both in real-world outcome studies,^10^ as well as in antibody reactivity studies, though the latter included much smaller cohorts.^34–36^

Interestingly, when examining younger versus older age groups, we found that ages 5-11 exhibited no significant waning of naturally acquired protection throughout the outcome period, whereas waning protection in the 12-18 age group was more prominent, but still mild. This finding is consistent with previous observations that pointed to lower reinfection rates in children compared to adults,^14,15^ and in younger children in particular.^37^ It has been demonstrated that children have a different profile of immune response after a SARS-CoV-2 infection compared to adults,^38^ though the biological paradigm that sufficiently explains this longer lasting protection requires further investigations.

Our analysis is subject to several limitations. First, the outcome period does not include Omicron-dominant months. The reason is that on January 2022 a major policy change took place in Israel, whereby PCR tests were not readily available for persons under 60 years old.^39^ Concomitantly, most rapid antigen tests were at-home tests, thus not reported back to our centralized database. Therefore, attempting to estimate infections during that time would lead to gross bias, underestimating infections. Thus, the protection of a previous infection against other novel strains, including the Omicron variant, cannot be directly inferred.

An inherent limitation to real-world observational studies of COVID-19 pertains to the lack of pre-defined PCR testing protocols. This could yield potential biases relating to test-taking and healthcare seeking behaviors, a matter intricately discussed in previous studies.^23,24,26,40^ The test-negative design aims to somewhat mitigate this potential bias,^32^ whereas those untested are not assumed to be COVID-free. Furthermore, the fact that our secondary design (Poisson regression), also prevalent in COVID-19 studies,^41^ yielded similar outcomes, reinforces our confidence in the results, while it also supports previous discussions highlighting the advantage of the test negative design in adjusting for changes in testing volume.^23,29,42^ Moreover, the secondary analysis included matching for previous PCR tests taken throughout the pandemic, as a proxy for healthcare seeking behavior.^30,40^ Additionally, in order to address potential differences between the groups, especially in terms of exposure (as their medical histories were relatively comparable), we chose a matched design by GSA and week of testing, specifically important due to time-varying incidence rates between geographical areas. In the corresponding Poisson analysis, we included adjustment for attack rates, calculated for each small residential area, represented by affiliation to one’s local care facility.

Lastly, only a small number of COVID-19 related hospitalizations were recorded, and no deaths, a statistical analysis of severe disease was not performed. As we reported, most hospitalizations were incurred by unvaccinated and previously uninfected individuals.

In conclusion, this study suggests that children and adolescents who were previously infected with SARS-CoV-2 remain protected against reinfection to a high degree for up to 18 months. In light of these findings, alongside evidence pointing to the fact that the majority of this population had already been infected, policy decision makers should consider when and if convalescent children and adolescents should be vaccinated. Nonetheless, further research is needed to examine naturally acquired immunity against emerging variants, including the Omicron.

## Supplementary Tables and Figures

**Figure S1.**
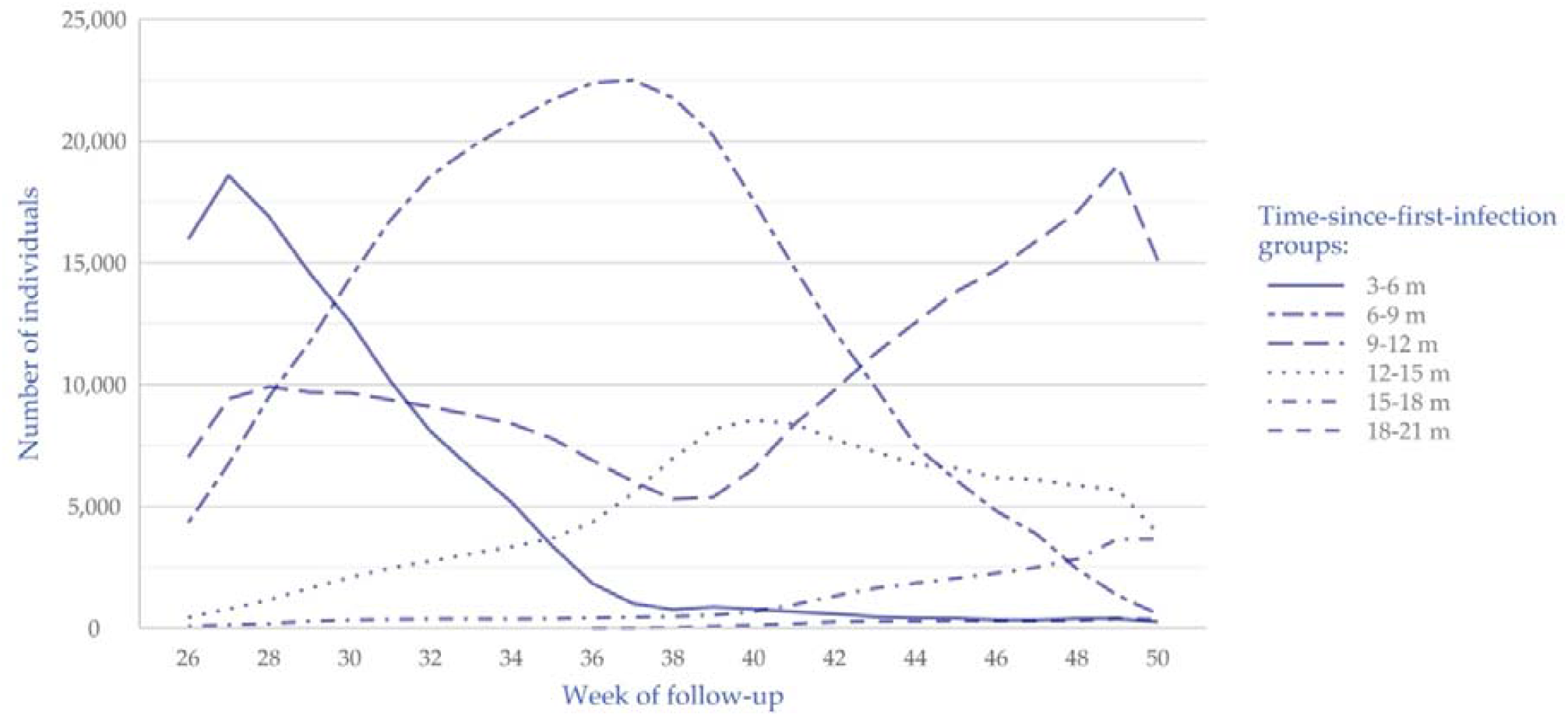
Dynamics of cohorts in the secondary analysis. The different graphs represent the number of eligible individuals in each cohort, throughout the follow-up period. The dynamics is explained by individuals exiting and entering groups, determined by to their immunity status (time-since-infection) on each follow-up day.

**Figure S2.**
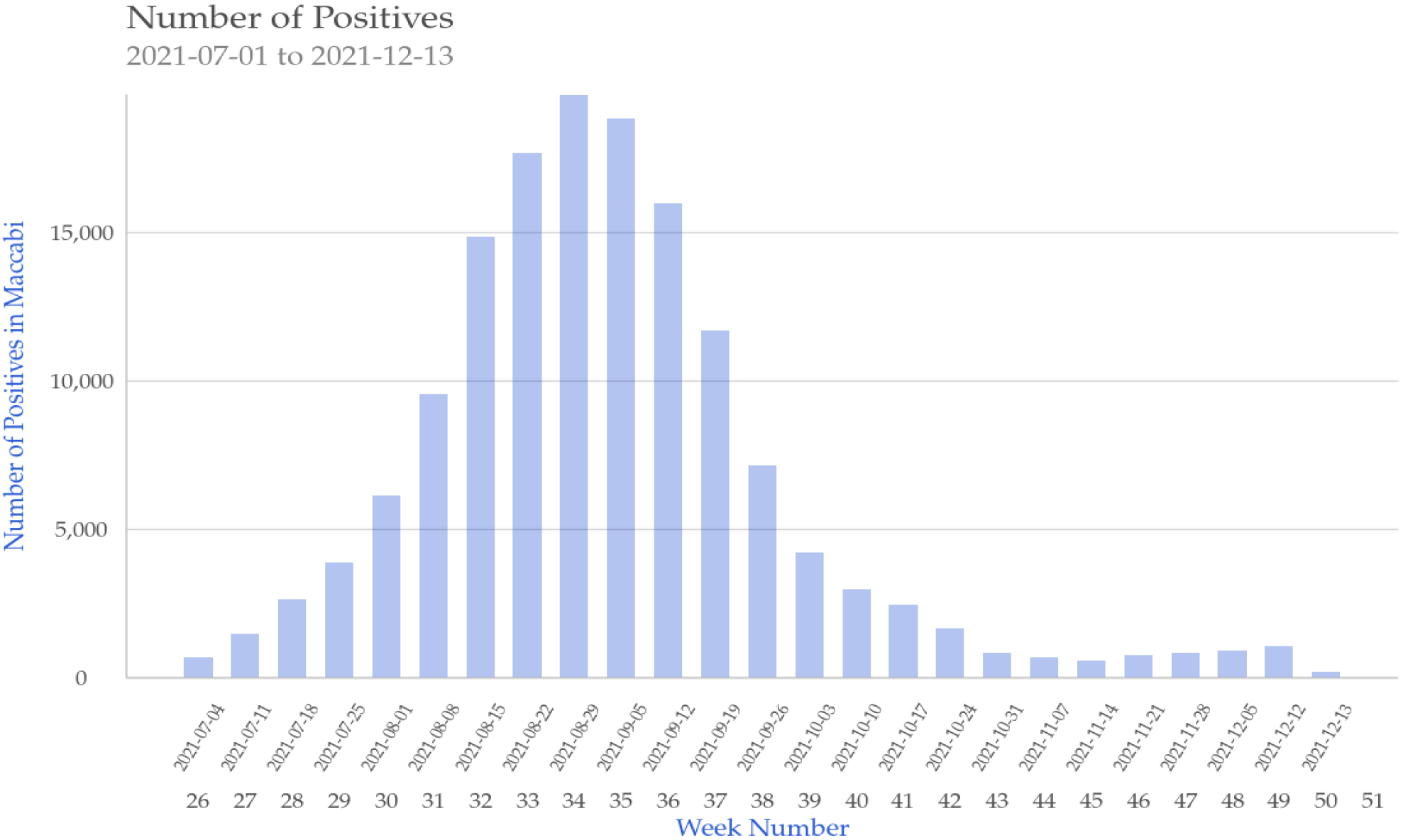
Weekly SARS-CoV-2 confirmed infections in the during the outcome period.

**Figure S3.**
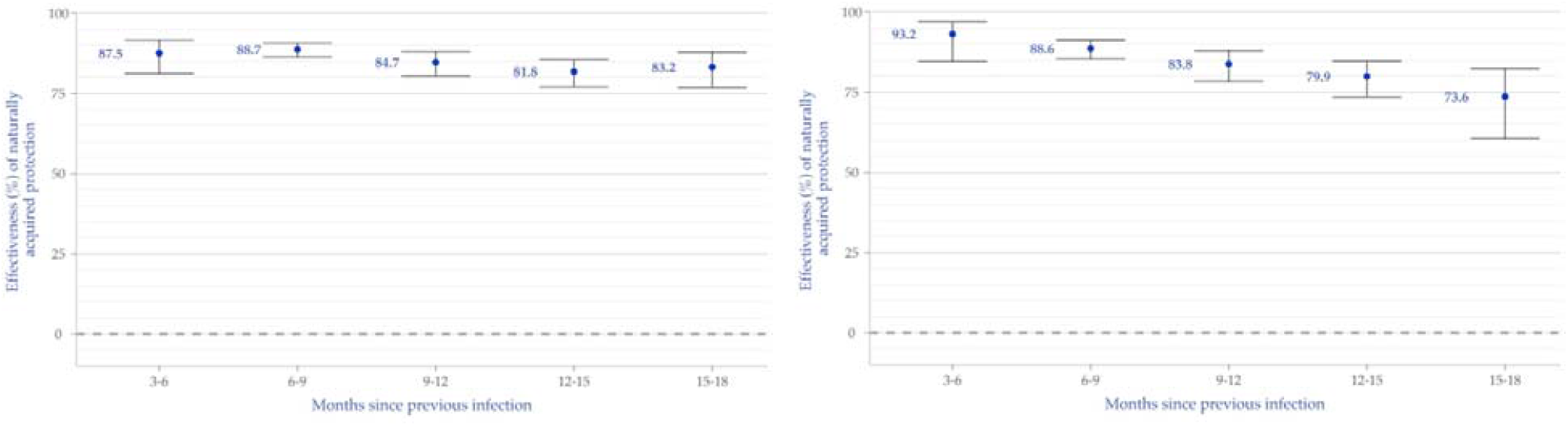
Additional analysis by age groups; reduction in the odds of testing positive for SARS-CoV-2 among children and adolescents who were previously infected compared to SARS-CoV-2 naïve individuals, by time since previous infection, based on the primary test-negative analysis. Bars represent 95% confidence intervals. Left panel: ages 5-11, right panel: ages 12-18

**Figure S4.**
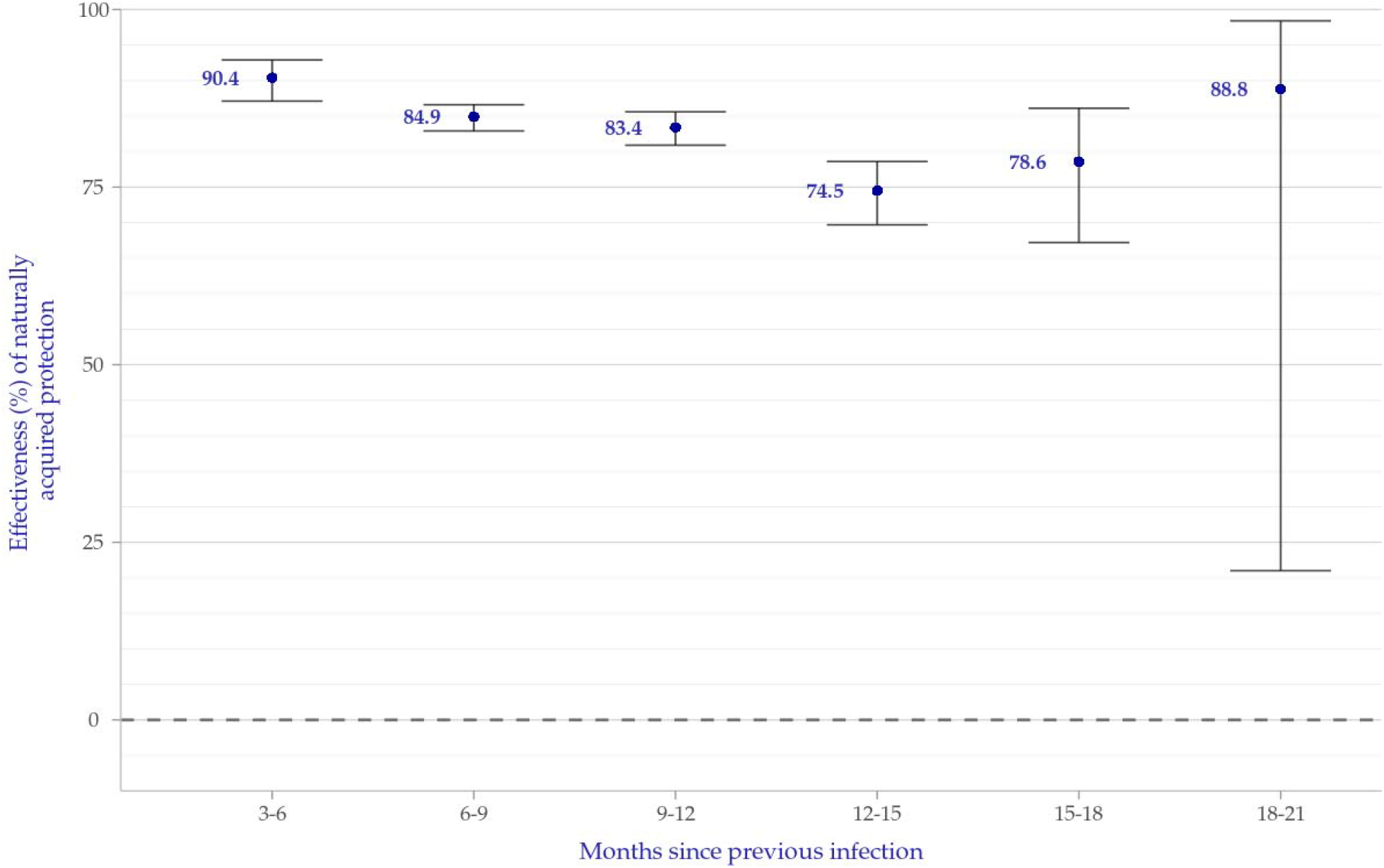
Effectiveness of a previous-infection against re-infection, calculated as 100%*[1-(Rate Ratio)] for each of the 3-month-interval-since-infection categories using a Poisson regression model. Bars represent 95% confidence intervals.

**Figure S5.**
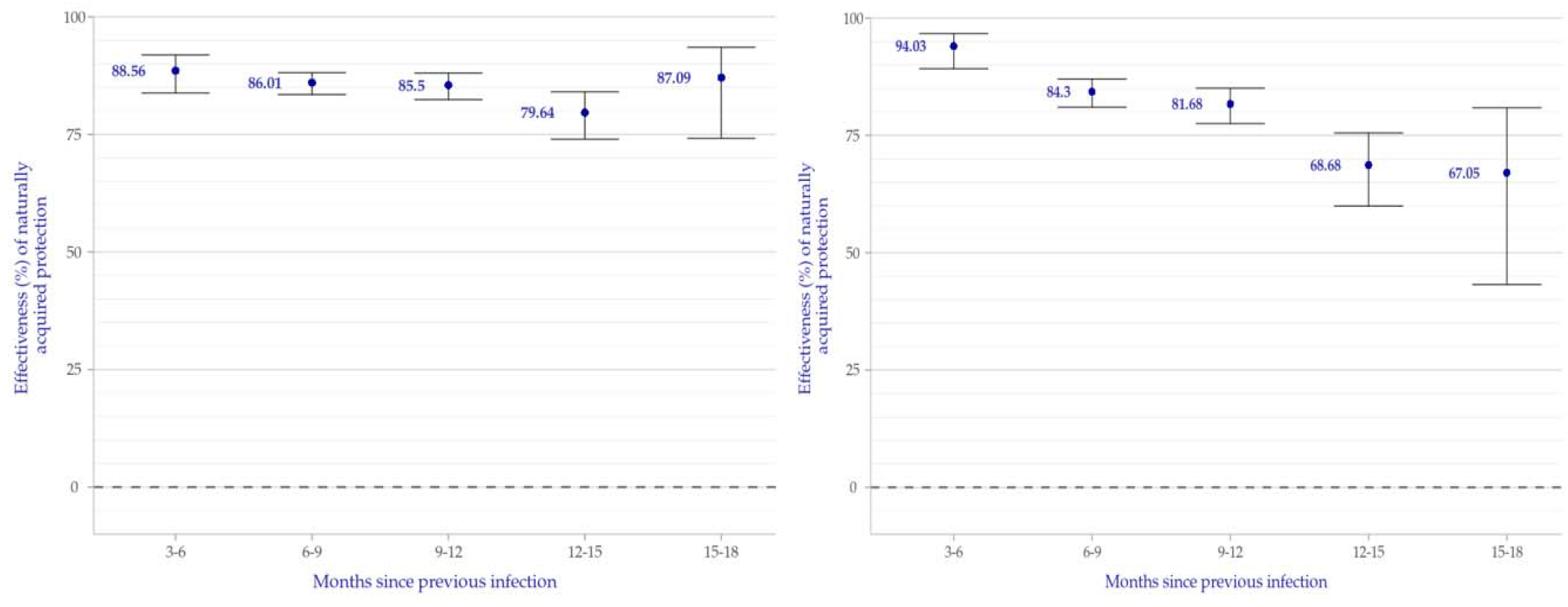
Additional analysis by age groups: effectiveness of a previous-infection against re-infection, calculated as 100%*[1-(Rate Ratio)] for each of the 3-month-interval-since-infection categories using a Poisson regression model. Bars represent 95% confidence intervals. Left panel: ages 5-11, right panel: ages 12-18

**Figure S6.**
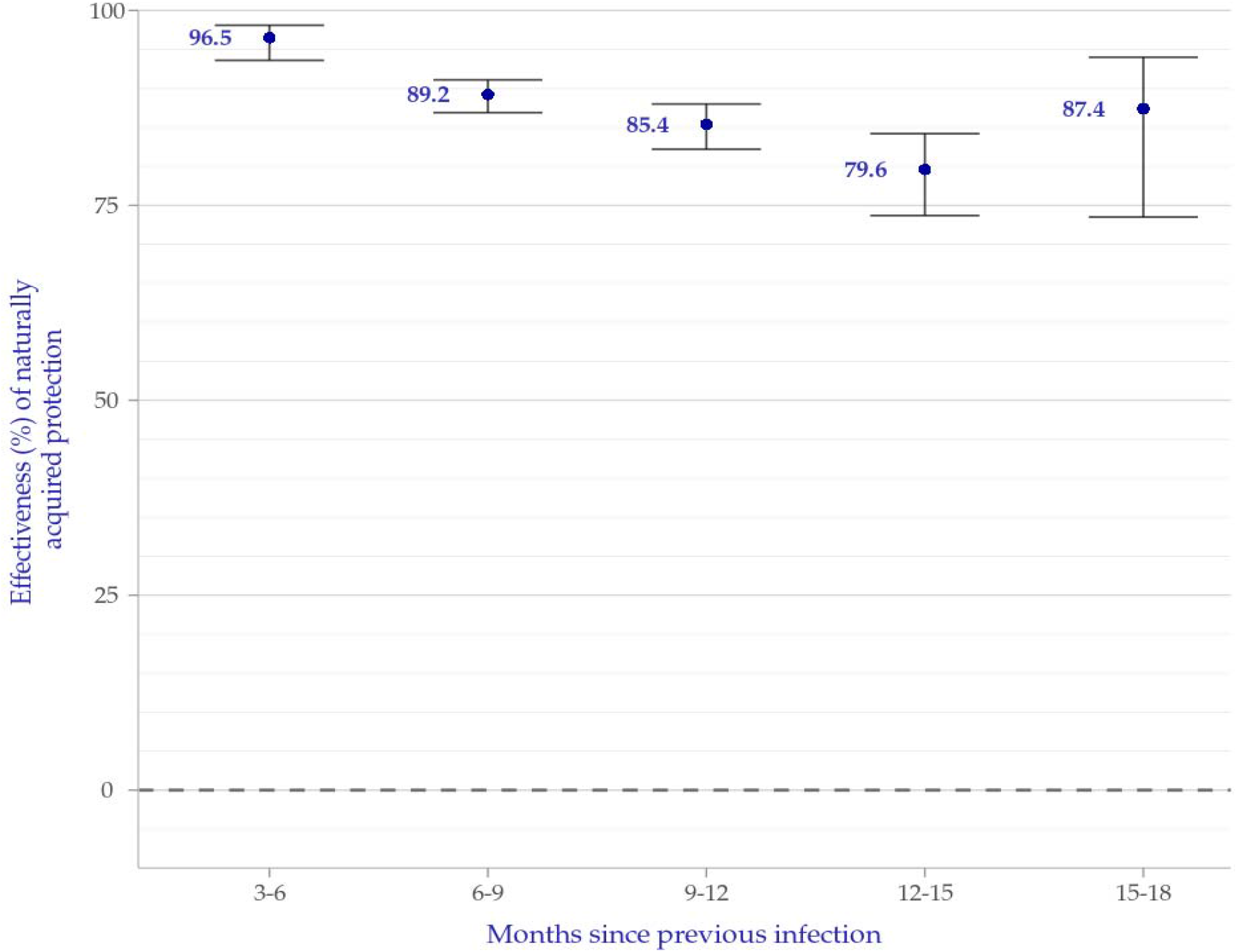
Effectiveness of a previous-infection against a symptomatic re-infection (COVID-19), calculated as 100%*[1-(Rate Ratio)] for each of the 3-month-interval-since-infection categories using a Poisson regression model. Bars represent 95% confidence intervals.

**Table S1.** Characteristics of previously infected participants, 5-18 years old, who were tested between July 1 and December 13, 2021, by groups. As group membership is dynamic, and participants can contribute person-days to multiple groups, distribution is presented as percentage of contributed person-days at risk rather than number or percentage of individuals.

|  | 3-6 months<br>after<br>infection | 6-9 months<br>after<br>infection | 9-12 months<br>after<br>infection | 12-15 months<br>after<br>infection | 15-18 months<br>after<br>infection | 18-21 months<br>after<br>infection |
| --- | --- | --- | --- | --- | --- | --- |
| Number of person-days at risk | 818065 | 2113613 | 1716632 | 817726 | 184644 | 20806 |
| *Characteristics – in % person-days at risk* |  |  |  |  |  |  |
| **Age distribution** |  |  |  |  |  |  |
| 5-11 | 61.4 | 66.3 | 66.8 | 66.2 | 66.4 | 61.8 |
| 12-18 | 38.6 | 33.7 | 33.2 | 33.8 | 33.6 | 38.2 |
| **Sex** |  |  |  |  |  |  |
| Male | 50.8 | 51.4 | 52.7 | 53.0 | 51.4 | 49.8 |
| Female | 49.2 | 48.6 | 47.3 | 47.0 | 48.6 | 50.2 |
| **SES** |  |  |  |  |  |  |
| Low (1-3) | 16.9 | 21.7 | 25.4 | 26.8 | 31.2 | 43.9 |
| Medium (4-6) | 50.4 | 50.9 | 49.6 | 48.6 | 46.3 | 41.6 |
| High (7-10) | 32.7 | 27.4 | 25.0 | 24.5 | 22.5 | 14.5 |
| Missing | 0.04 | 0.03 | 0.03 | 0.04 | 0.02 | 0 |
| **Comorbidities** |  |  |  |  |  |  |
| Immunosuppression | 0.01 | 0.003 | 0 | 0 | 0 | 0 |
| DM | 0.16 | 0.14 | 0.15 | 0.13 | 0.09 | 0 |
| Cancer | 0.15 | 0.16 | 0.18 | 0.15 | 0.12 | 0 |
| Obesity (BMI ≥30) | 1.79 | 1.52 | 1.56 | 1.79 | 1.79 | 2.28 |
| IBD | 0.12 | 0.12 | 0.09 | 0.04 | 0.05 | 0.07 |
| CVD | 1.36 | 1.34 | 1.26 | 1.19 | 1.26 | 2.38 |
| CKD | 0.01 | 0.004 | 0.001 | 0 | 0 | 0 |

SES – Socioeconomic status on a scale from 1 (lowest) to 10; DM – Diabetes Mellitus; CKD – Chronic Kidney Disease; COPD – Chronic Obstructive Pulmonary Disease; BMI – Body Mass Index.

**Table S2.** Left column represents the adjusted effectiveness 100%*[1-Rate Ratio] based on a Poisson regression analysis for each time-since-first-infection group relative to SARS-CoV-2 naïve individuals. Right column demonstrates the adjusted incidence rates of SARS-CoV-2 infection per 100,000 person days for each time-since-infection group. For each column, the 95% confidence intervals are presented in brackets.

| Groups of months-since-previous-infection | Outcome: SARS-CoV-2 infection |  | Outcome: SARS-CoV-2 symptomatic infection |  |
| --- | --- | --- | --- | --- |
|  | VE(1-RR) of reinfections compared to SARS-CoV-2 naïve persons and 95% C.I. | Adjusted incidence rates of SARS-CoV-2 infection per 100,000 person days | VE(1-RR) of symptomatic reinfections (COVID-19) compared to SARS-CoV-2 naïve persons and 95% C.I. | Adjusted incidence rates of COVID-19 per 100,000 person days |
| Previously Uninfected |  | 75.78(74.99, 76.52) |  | 36.5(35.97, 37.02) |
| 3-6 | 0.9(0.87, 0.93) | 7.22(5.22, 9.59) | 0.97(0.94, 0.98) | 1.26(0.51, 2.12) |
| 6-9 | 0.85(0.83, 0.87) | 11.47(9.94, 12.95) | 0.89(0.87, 0.91) | 3.92(3.2, 4.7) |
| 9-12 | 0.83(0.81, 0.86) | 12.56(10.83, 14.29) | 0.85(0.82, 0.88) | 5.38(4.36, 6.53) |
| 12-15 | 0.75(0.7, 0.79) | 19.29(16.09, 22.82) | 0.8(0.74, 0.84) | 7.49(5.6, 9.51) |
| 15-18 | 0.79(0.67, 0.86) | 16.07(9.32, 24.09) | 0.87(0.74, 0.94) | 4.63(1.33, 8.51) |
| 18-21 | 0.89(0.21, 0.98) | 8.42(0, 25.93) |  |  |

## References

1. Krammer, F. A correlate of protection for SARS-CoV-2 vaccines is urgently needed. Nat. Med. 2021 277 27, 1147–1148 (2021).

2. Iwasaki, A. What reinfections mean for COVID-19. Lancet Infect. Dis. 21, 3–5 (2021).

3. Tomassini, S. et al. Setting the criteria for SARS-CoV-2 reinfection–six possible cases. J. Infect. 82, 282.

4. Coronavirus Disease 2019 (COVID-19) 2021 Case Definition | CDC. https://ndc.services.cdc.gov/case-definitions/coronavirus-disease-2019-2021/ (2021).

5. Perez, G. et al. A 1 to 1000 SARS-CoV-2 reinfection proportion in members of a large healthcare provider in Israel: a preliminary report. MedRxiv http://medrxiv.org/content/early/2021/03/08/2021.03.06.21253051.abstract (2021) doi:10.1101/2021.03.06.21253051.

6. Lumley, S. F. et al. Antibody Status and Incidence of SARS-CoV-2 Infection in Health Care Workers. N. Engl. J. Med. 384, 533–540 (2021).

7. Kim, P., Gordon, S. M., Sheehan, M. M. & Rothberg, M. B. Duration of Severe Acute Respiratory Syndrome Coronavirus 2 Natural Immunity and Protection Against the Delta Variant: A Retrospective Cohort Study. Clin. Infect. Dis. ciab999 (2021) doi:10.1093/CID/CIAB999.

8. Hansen, C. H., Michlmayr, D., Gubbels, S. M., Mølbak, K. & Ethelberg, S. Assessment of protection against reinfection with SARS-CoV-2 among 4 million PCR-tested individuals in Denmark in 2020: a population-level observational study. Lancet 397, 1204–1212 (2021).

9. Abu-Raddad, L. J. et al. Relative infectiousness of SARS-CoV-2 vaccine breakthrough infections, reinfections, and primary infections. Nat. Commun. 2022 131 13, 1–11 (2022).

10. Gazit, S. et al. SARS-CoV-2 Naturally Acquired Immunity vs. Vaccine-induced Immunity, Reinfections versus Breakthrough Infections: a Retrospective Cohort Study. Clin. Infect. Dis. (2022) doi:10.1093/CID/CIAC262.

11. Ministry of Health’s Position Regarding the Expansion of the Vaccination Operation to Ages 12-16 Years | Ministry of Health. https://www.gov.il/en/departments/news/02062021-01.

12. Food and Drug Administration (FDA). Coronavirus (COVID-19) Update: FDA Authorizes Pfizer-BioNTech COVID-19 Vaccine for Emergency Use in Adolescents in Another Important Action in Fight Against Pandemic. Food and Drug Administration (FDA) https://www.fda.gov/news-events/press-announcements/coronavirus-covid-19-update-fda-authorizes-pfizer-biontech-covid-19-vaccine-emergency-use (2021).

13. First COVID-19 vaccine approved for children aged 12 to 15 in EU | European Medicines Agency. https://www.ema.europa.eu/en/news/first-covid-19-vaccine-approved-children-aged-12-15-eu.

14. Slezak, J. et al. Rate and severity of suspected SARS-Cov-2 reinfection in a cohort of PCR-positive COVID-19 patients. Clin. Microbiol. Infect. 27, 1860.e7-1860.e10 (2021).

15. Mensah, A. A. et al. Risk of SARS-CoV-2 reinfections in children: a prospective national surveillance study between January, 2020, and July, 2021, in England. Lancet Child Adolesc. Heal. 6, 384–392 (2022).

16. Reinfection. Centers for Disease Control and Prevention (CDC) https://www.cdc.gov/coronavirus/2019-ncov/php/reinfection.html (2020).

17. Shalev, V. et al. The use of an automated patient registry to manage and monitor cardiovascular conditions and related outcomes in a large health organization. Int. J. Cardiol. 152, 345–349 (2011).

18. Weitzman, D., Chodick, G., Shalev, V., Grossman, C. & Grossman, E. Prevalence and factors associated with resistant hypertension in a large health maintenance organization in Israel. Hypertension 64, 501–507 (2014).

19. Chodick, G., Heymann, A. D., Shalev, V. & Kookia, E. The epidemiology of diabetes in a large Israeli HMO. Eur. J. Epidemiol. 18, 1143–1146 (2003).

20. Coresh, J. et al. Decline in estimated glomerular filtration rate and subsequent risk of end-stage renal disease and mortality. JAMA - J. Am. Med. Assoc. 311, 2518–2531 (2014).

21. Israel National Cancer Registry. Israel Center for Disease Control https://www.health.gov.il/English/MinistryUnits/HealthDivision/Icdc/Icr/Pages/default.aspx.

22. SARS-CoV-2 variants in analyzed sequences, Israel. https://ourworldindata.org/grapher/covid-variants-area?country=~ISR.

23. Patalon, T. et al. Odds of Testing Positive for SARS-CoV-2 Following Receipt of 3 vs 2 Doses of the BNT162b2 mRNA Vaccine. JAMA Intern. Med. (2021) doi:10.1001/JAMAINTERNMED.2021.7382.

24. Mizrahi, B. et al. Correlation of SARS-CoV-2-breakthrough infections to time-from-vaccine. Nat. Commun. 12, 1–5 (2021).

25. Chemaitelly, H. et al. Waning of BNT162b2 Vaccine Protection against SARS-CoV-2 Infection in Qatar. N. Engl. J. Med. 385, e83 (2021).

26. Patalon, T. et al. Waning Effectiveness of the Third Dose of the BNT162b2 mRNA COVID-19 Vaccine. medRxiv 2022.02.25.22271494 (2022) doi:10.1101/2022.02.25.22271494.

27. Gazit, S. et al. Relative Effectiveness of Four Doses Compared to Three Dose of the BNT162b2 Vaccine in Israel. medRxiv 2022.03.24.22272835 (2022) doi:10.1101/2022.03.24.22272835.

28. Dean, N. E., Hogan, J. W. & Schnitzer, M. E. Covid-19 Vaccine Effectiveness and the Test-Negative Design. https://doi.org/10.1056/NEJMe2113151 385, p1431–1433 (2021).

29. Abu-Raddad, L. J., Chemaitelly, H. & Butt, A. A. Effectiveness of the BNT162b2 Covid-19 Vaccine against the B.1.1.7 and B.1.351 Variants. N. Engl. J. Med. 385, 187–189 (2021).

30. Mizrahi, B. et al. Correlation of SARS-CoV-2-breakthrough infections to time-from-vaccine. Nat. Commun. 12, 1–5 (2021).

31. Goldberg, Y. et al. Waning Immunity after the BNT162b2 Vaccine in Israel. N. Engl. J. Med. 385, e85 (2021).

32. Gazit, S. et al. Short term, relative effectiveness of four doses versus three doses of BNT162b2 vaccine in people aged 60 years and older in Israel: retrospective, test negative, case-control study. BMJ 377, e071113 (2022).

33. Clarke, K. E. N. et al. Morbidity and Mortality Weekly Report Seroprevalence of Infection-Induced SARS-CoV-2 Antibodies-United States, September 2021-February 2022. https://www.medrxiv.org/content/10.1101/2022.04.18.22271936v1?rss doi:10.1101/2022.04.18.22271936v1?rss.

34. Wang, Z. et al. Naturally enhanced neutralizing breadth against SARS-CoV-2 one year after infection. Nature 595, 426–431 (2021).

35. Gallais, F. et al. Evolution of antibody responses up to 13 months after SARS-CoV-2 infection and risk of reinfection. EBioMedicine 71, 103561 (2021).

36. Niemi, M. E. K. et al. Mapping the human genetic architecture of COVID-19. Nat. 2021 6007889 600, 472–477 (2021).

37. Kubale, J. et al. Burden of SARS-CoV-2 and protection from symptomatic second infection in children. medRxiv 2022.01.03.22268684 (2022) doi:10.1101/2022.01.03.22268684.

38. Dowell, A. C. et al. Children develop robust and sustained cross-reactive spike-specific immune responses to SARS-CoV-2 infection. Nat. Immunol. 2021 231 23, 40–49 (2021).

39. Changes in the framework of testing: PCRs to 60 and older and at risk populations. Vaccinated - to home tests. YNET https://www.ynet.co.il/news/article/rjthxkq2t#autoplay.

40. Bar-On, Y. M. et al. Protection of BNT162b2 Vaccine Booster against Covid-19 in Israel. N. Engl. J. Med. 385, 1393–1400 (2021).

41. Goldberg, Y. et al. Protection and waning of natural and hybrid COVID-19 immunity. medRxiv 2021.12.04.21267114 (2021) doi:10.1101/2021.12.04.21267114.

42. Dean, N. E., Hogan, J. W. & Schnitzer, M. E. Covid-19 Vaccine Effectiveness and the Test-Negative Design. https://doi.org/10.1056/NEJMe2113151 385, 1431–1433 (2021).

